# Associations between health literacy and sociodemographic factors: A cross-sectional study in Malaysia utilising the HLS-M-Q18

**DOI:** 10.1101/2021.02.19.21252088

**Authors:** Arina Anis Azlan, Mohammad Rezal Hamzah, Tham Jen Sern, Suffian Hadi Ayub, Abdul Latiff Ahmad, Emma Mohamad

**Affiliations:** Centre for Research in Media and Communication, Faculty of Social Sciences and Humanities, Universiti Kebangsaan Malaysia, Bangi, Selangor, Malaysia; HEALTHCOMM UKM x UNICEF C4D Centre, Faculty of Social Sciences and Humanities, Universiti Kebangsaan Malaysia, Bangi, Selangor, Malaysia; Centre of Excellence for Social Innovations and Sustainability, Faculty of Applied and Human Sciences, Universiti Malaysia Perlis, Kangar, Perlis, Malaysia; Department of Communication, Faculty of Modern Languages and Communication, Universiti Putra Malaysia, Seri Kembangan, Selangor, Malaysia; Faculty of Communication and Media Studies, Universiti Teknologi MARA, Shah Alam, Selangor, Malaysia

## Abstract

Health literacy is progressively seen as an indicator to describe a nation’s health status. To improve health literacy, countries need to address health inequalities by examining different social demographic factors across the population. This assessment is crucial to identify and evaluate strengths and limitations of a country in addressing health issues. By addressing these health inequalities, a country would be better informed to take necessary steps to improve the nation’s health literacy. This study examines health literacy levels in Malaysia and analyses socio-demographic factors that are associated with health literacy. A cross-sectional survey was carried out using the HLS-M-Q18 instrument which was validated for the Malaysian population. Multi-stage random sampling strategy was used in this study utilising several sampling techniques including quota sampling, cluster sampling and simple random sampling to allow random data collection. A total of 855 respondents were sampled. Results found significant associations between health literacy and age, health status and health problems. Findings also suggest that lower health literacy levels were found to be associated with the younger generation. The findings of this study have provided baseline data of the health literacy of Malaysians and provide evidence toward potential areas of intervention.

## 1. Introduction

Worldwide interest in studying health literacy is increasing as health promoters and practitioners recognise its significance in reducing illness [1] and improving quality of life [2]. The benefits of health literacy extend beyond individual health care to include effective disease prevention in society, as well as improving health promotion in general. Health literacy is a concept that extends beyond health education. It addresses social and environmental factors that influences individual ability to engage with health information to make informed decisions and to utilise health services to benefit them and their surroundings.

Studies have emphasised a variety of health literacy benefits to society [3] and have reported risks of populations with low health literacy [4]. Higher health literacy has also been associated with positive health outcomes [5], health behaviours [5,6] and lower health cost [7]. It is plausible to expect that the way forward to better global health management is through improvement of society’s health literacy levels.

As awareness on the importance of health literacy increases, more instruments are being developed to accurately measure health literacy rates among general populations, as well as in specific groups. A 2017 systematic review revealed 36 instruments used to measure health literacy [8] while another systematic review in 2018 reported 29 health literacy instruments used on children and adolescents [9].

In Malaysia, several tools have been utilised to measure health literacy [10], however an extensive study across six Asian countries including Malaysia has demonstrated that the European Health Literacy Survey Questionnaire (HLS-EU-Q47) instrument has good construct validity, satisfactory goodness-of-fit to the three health literacy domains (health care, disease prevention and health promotion), as well as high internal consistency and satisfactory item-scale convergent validity [11]. This suggests that the HLS-EU-Q47 instrument may serve as a good indicator to measure health literacy for the Malaysian society. A recent study has further compressed the HLS-EU-Q47 into a short scale instrument, the HLS-M-Q18, consisting 18 items to accommodate for the Malaysian National Health Morbidity Survey in 2019 [12]. The HLS-EU-Q47 and HLS-M-Q18 instruments operationally defined health literacy as the ability to access, understand, appraise and apply health information across three domains; health care, disease prevention and health promotion. This model was conceptualised by an extensive systematic review which integrated health literacy definitions and models with the aim to develop a comprehensive evidence-based dimensions of health literacy.

The objectives of this study are to (1) measure society’s health literacy and (2) observe socio-demographic factors that are associated with health literacy in Malaysia. In order to improve health inequalities in the community, an assessment of individual health literacy is crucial to identify and evaluate the strengths and limitations in addressing health issues in a diverse society [13]. In order for health care providers and policy makers to respond efficiently, they need to understand the diverse factors that affect health literacy before facilitating access to health information, providing services and devising health intervention that does not discriminate against health literacy limitations [14].

## 2. Methods

### Study design

A nationally representative cross-sectional survey was employed to address the research objectives. The survey was administered by well-trained enumerators who were also staff working for the Ministry of Health Malaysia. Three states were selected (Selangor, Kuala Lumpur and Sarawak) to represent the distribution of multiple ethnicities, as well as the distribution of urban and rural areas. The selection of areas was made based on referral and advice by the District of Jurisdiction Malaysia, Rural Master Plan Malaysia, and previous literature [15].

### Ethical approval

The National Medical Ethics Committee Malaysia under the Ministry of Health Malaysia approved our study protocol, procedures, information sheet and consent statement (NMRR-18-1320/41882). All respondents were above 18 years old and therefore involved no minors. All respondents also signed a written consent form clearly stating their rights and nature of participation in the study before being asked to answer the questionnaire. The confidentiality of the information and privacy of the respondents were protected throughout the study.

### Recruitment procedure

Multi-stage random sampling was used in this study. In detail, there were three stages involved, utilising several sampling techniques (quota sampling, cluster sampling and simple random sampling) to allow random data collection. The three stages are illustrated in Figure 1.

**Fig 1.** The multi-stage random sampling procedure.

In stage 1, quota sampling based on ethnicities and urban or rural distribution were used to select three Malaysian states. Ethnic distribution should be a standard in sampling multiracial populations to ensure inclusivity of the sample [16]. States from both Peninsular Malaysia and Borneo were selected to represent the diverse ethnicities in Malaysia. For the purpose of urban and rural distribution, Kuala Lumpur and Sarawak were selected to represent the urban and rural areas respectively. This is justified as Kuala Lumpur has the highest urban population while Sarawak has the highest rural population in Malaysia. In selecting the state of Sarawak, a more balanced representation of the minority ethnic groups could be obtained (i.e., Bumiputera). Selangor represents both urban and rural areas and has a balanced ratio in ethnic group distribution.

In stage 2, cluster sampling was utilised to determine districts of choice. District sampling for Selangor was determined based on the demographic distribution list published by the Selangor Economic Development Unit, as well as extant literature [17]. For selection of districts in Kuala Lumpur, researchers used data provided by the Department of Information, Ministry of Communications and Multimedia Malaysia; and for Sarawak, the selection of districts was guided by data provided by the State Director of the Fire and Rescue Department. The definitions of rural and urban were determined by the National Department of Statistics and The Rural Master Plan, published by the Ministry of Rural Development Malaysia [18].

In stage 3, respondents were selected using a simple random sampling technique based on several criteria (i.e., Malaysian, aged 18 and above, resident in the chosen state, able to make health decisions for themselves). The respondent recruitment method used in this study mirrors the method and protocol criteria used by the Asian Health Literacy Consortium and previous literature [19]. If there were no eligible respondents in the household who met the selection criteria, household members were thanked for their time and the enumerator approached the next selected household. Only one respondent from any given household was interviewed, in which the eldest household member would be chosen if there was more than one household member who met the respondent selection criteria.

The researchers made the decision to prioritise an inclusive Malaysian sample based on ethnicity and urban/rural strata due to constraints in resources. This was to ensure that the smaller groups were adequately represented in the sample. The list of states, ethnicities and urban/rural distribution required for this study are as presented in Table 1.

**Table 1.** Sample distribution.

| State | Locality | Ethnicity | n | Area |
| --- | --- | --- | --- | --- |
| Selangor | Urban | Malay | 299 | Shah Alam |
| (6.298 mil =<br>58.2%)<br>N = 466 | (93.3%)<br>N = 435 | Chinese | 103 |  |
|  |  | Indian | 33 |  |
|  | Rural<br>(6.7%)<br>N = 31 | Malay | 21 | Hulu Langat |
|  |  | Chinese | 7 |  |
|  |  | Indian | 3 |  |
| Kuala Lumpur<br>(1.782 mil =<br>16.5%)<br>N = 132 | Urban<br>(100%)<br>N = 132 | Malay | 82 | Segamat and Lembah Pantai |
|  |  | Chinese | 38 |  |
|  |  | Indian | 12 |  |
| Sarawak<br>(2.741 mil =<br>25.3%)<br>N = 202 | Urban<br>(57.8%)<br>N = 117 | Bumiputera | 91 | Kuching |
|  |  | Chinese | 26 |  |
|  |  | Indian | 0 |  |
|  | Rural<br>(42.2%)<br>N = 85 | Bumiputera | 66 | Sarikei (Maradong) and Samarahan<br>(Simujan) |
|  |  | Chinese | 19 |  |
|  |  | Indian | 0 |  |

Note : Location of study is determined by population density and ethnic distribution

The cross-sectional survey was conducted between 25th June 2018 to 14th July 2018, involving 18 enumerators. All trained enumerators were working for the Ministry of Health Malaysia, wore the Ministry’s uniform and presented their identity cards to avoid misunderstanding and protect the interest of both researchers and respondents. Respondents took an average of 30–40 mins to complete the questionnaire. The target sample size was 470, determined by identifying the smallest acceptable size of a demographic subgroup with a ±5% margin of error and a confidence level of 95% [20-22]. The enumerators went from household to household within the selected areas and provided the self-reported questionnaire to be answered. A consent form was filled in and obtained from each respondent A total of 866 complete responses with no missing data were obtained and analysed.

### Study instrument

The survey instrument was adopted from the HLS-M-Q18 short version of health literacy questionnaire which was validated in a study [12]. The questionnaire contained three main sections: 1) demographics, which surveyed respondents’ socio-demographic information, including gender, age, race, marital status and income; 2) personal health information; 3) 18-item measure of health literacy. The questionnaire was constructed in the English and Malay languages. A backward-translation approach was used in translating the items between English and Malay, so as to ensure linguistic and conceptual equivalence [22]. Discrepancies between the two versions were rectified, and equivalence of measuring on all items was ensured through consultation with bilingual researchers.

Personal health information was measured by three items. First, respondents were asked to rate their health condition from “bad” coded as “1” to “good” coded as “2”. The second item asked respondents to identify if they suffered from long-term illness: “Do you have any long-term illness or health problems? Long-term illness means problems which have lasted, or you expect to last, 6 months or more”. Two answer options were provided (1 = No and 1 = Yes, one or more than one). The third item asked respondents to identify their frequency of involvement in physical activities such as lifting and carrying heavy objects, hoeing, mopping the floor or exercise (such as cycling, walking or jogging) for at least 10 minutes in the past 7 days.

To measure respondents’ health literacy, 18 items were adopted from a validated Malaysian version of the HLS-EU-Q47 [12]. Respondents were asked to identify the level of difficulty, ranging from 1 = very difficult to 4 = very easy. An index was created based on the above 18 items (S1 Appendix).

Demographic variables were controlled to reduce confounding effects. These variables included age, year of birth (1950 to 1965 for Baby Boomers, 1966 – 1976 for Generation X, 1977 to 1994 for Generation Y and 1995 to 2012 for Generation Z), gender (0 = female, 1 = male), race (1 = Malay/Bumiputera, 2 = Chinese, 3 = Indian), marital status (1 = not married, 2 = married, 3 = separated/divorced, 4 = widowed), and monthly household income (ranging from 1 = below RM3,000, including no income, 2 = RM3,001 to RM9,000 and 3 = RM9,001 or more).

### Statistical analysis

For this study, the collected data were analysed using the Statistical Package for the Social Sciences (SPSS), version 26. Descriptive analysis focused on frequencies and percentages. Logistic regression tests, using the Forward: LR method were conducted to examine the relationships between control variables and personal health information and health literacy. For this analysis, the levels of health literacy were re-coded to 1 = limited (inadequate and problematic) and 2 = adequate (sufficient and excellent). Odd ratios (OR), 95% confidence intervals (CI), and their corresponding *P* values are reported as indicators of the magnitude and statistical significance of associations.

## 3. Results

### Demographic characteristics

A total of 866 respondents from different demographic segments and backgrounds participated in this study. The demographics were broadly representative of the Malaysian population with slightly fewer male participants at 34.9 % male and 65.1% female. Almost 70% of the study participants were from the young generation (Z and Y). Table 1 shows the distribution of respondents according to selected demographics. The majority of the respondents were female, Malay, generations Y and Z, not married, and had low income levels.

**Table 1.** Distribution of respondent characteristics and health literacy levels using HLS-M-Q18 (N=866).

|  | n (%) | Health Literacy level n (%) |  |  |  |
| --- | --- | --- | --- | --- | --- |
|  |  | Inadequate | Problematic | Sufficient | Excellent |
| **Respondents** | 866 (100) | 154 (17.8) | 348 (40.2) | 284 (32.8) | 79 (9.1) |
| **Age (mean)** | 866 (33.6) | 33.1 | 33.8 | 33.5 | 33.8 |
| Gen Z<br>(1995 – 2012) | 211 (24.4) | 33 (15.6) | 75 (35.5) | 78 (37.0) | 25 (11.8) |
| Gen Y<br>(1977 – 1994) | 377 (43.6) | 73 (19.4) | 162 (43.0) | 114 (30.2) | 28 (7.4) |
| Gen X<br>(1966 – 1976) | 184 (21.3) | 34 (18.5) | 77 (41.8) | 58 (31.5) | 15 (8.2) |
| Baby Boomers<br>(1950 – 1965) | 93 (9.8) | 13 (14.0) | 34 (36.6) | 35 (37.6) | 11 (11.8) |
| **Gender** |  |  |  |  |  |
| Male | 303 (35) | 65 (21.5) | 109 (36.0) | 105 (34.7) | 24 (7.9) |
| Female | 563 (65) | 89 (15.8) | 239 (42.5) | 180 (32.0) | 55 (9.8) |
| **Race** |  |  |  |  |  |
| Malay | 470 (54.3) | 68 (14.5) | 188 (40.0) | 170 (36.2) | 44 (9.4) |
| Chinese | 213 (24.6) | 46 (21.6) | 89 (41.8) | 59 (27.7) | 19 (8.9) |
| Indian | 65 (7.5) | 15 (23.1) | 21 (32.3) | 23 (35.4) | 6 (9.2) |
| Bumiputera | 115 (13.3) | 25 (21.7) | 48 (41.7) | 32 (27.8) | 10 (8.7) |
| **Marital status** |  |  |  |  |  |
| Not married | 429 (49.7) | 74 (17.2) | 163 (38.0) | 149 (34.7) | 43 (10.0) |
| Married | 394 (45.6) | 74 (18.8) | 162 (41.1) | 125 (31.7) | 33 (8.4) |
| Separated /<br>Divorced | 21 (2.4) | 4 (19.0) | 12 (57.1) | 4 (19.0) | 1 (4.8) |
| Widowed | 20 (2.3) | 2 (10.0) | 10 (50.0) | 6 (30.0) | 2 (10.0) |
| **Income** |  |  |  |  |  |
| Below<br>RM3,000<br>(including no<br>income) | 510 (59.4) | 88 (17.3) | 220 (43.1) | 162 (31.8) | 40 (7.8) |
| RM3,001 –<br>RM9,000 | 293 (34.1) | 58 (19.8) | 105 (35.8) | 99 (33.8) | 31 (10.6) |
| ≥RM9,001 | 55 (6.4) | 7 (12.7) | 20 (36.4) | 20 (36.4) | 8 (14.5) |
| **Exercise (days**<br>**a week)** |  |  |  |  |  |
| 0-2 days | 347(40.1) | 85 (24.5) | 130 (37.5) | 111 (32.0) | 21 (6.1) |
| More than 2<br>days a week | 519 (59.9) | 69 (13.3) | 218 (42.0) | 174 (33.5) | 58 (11.2) |
| **Health**<br>**Problems** |  |  |  |  |  |
| 1 and more<br>than 1 disease | 219 (25.3) | 42 (19.2) | 84 (38.4) | 76 (34.7) | 17 (7.8) |
| No disease | 646 (74.7) | 17.3 (17.3) | 40.9 (40.9) | 32.2 (32.2) | 9.6 (9.6) |
| **Health status** |  |  |  |  |  |
| Bad | 248 (28.7) | 70 (28.2) | 105 (42.3) | 61 (24.6) | 12 (4.8) |
| Good | 617 (71.3) | 84 (13.6) | 242 (39.2) | 224 (36.3) | 67(10.9) |

Over 28% of the respondents perceived their general health was bad, but over 70% perceived their health status to be very good or fairly healthy. Of the 866 respondents, 277 (17.8%) had inadequate health literacy, another 40.4% had marginal health literacy, 32.9% had adequate health literacy and 9.1% had excellent or very good health literacy. On the average, the results of the study show that the younger generation (aged 33.1.-33.8 years) crossed all levels of health literacy.

Several socio-demographic characteristics were associated with health literacy level and are shown in Table 2. Characteristics associated with health literacy level included health status, health problems, and age. The logistic regression model was statistically significant, χ2 (4) = 49.285, p < .000. The model explained 7.6% (Nagelkerke R2) of the variance. Hosmer and Lemeshow Test showed that the model is a good fit to the data as p=0.954 (>.05).

**Table 2.**
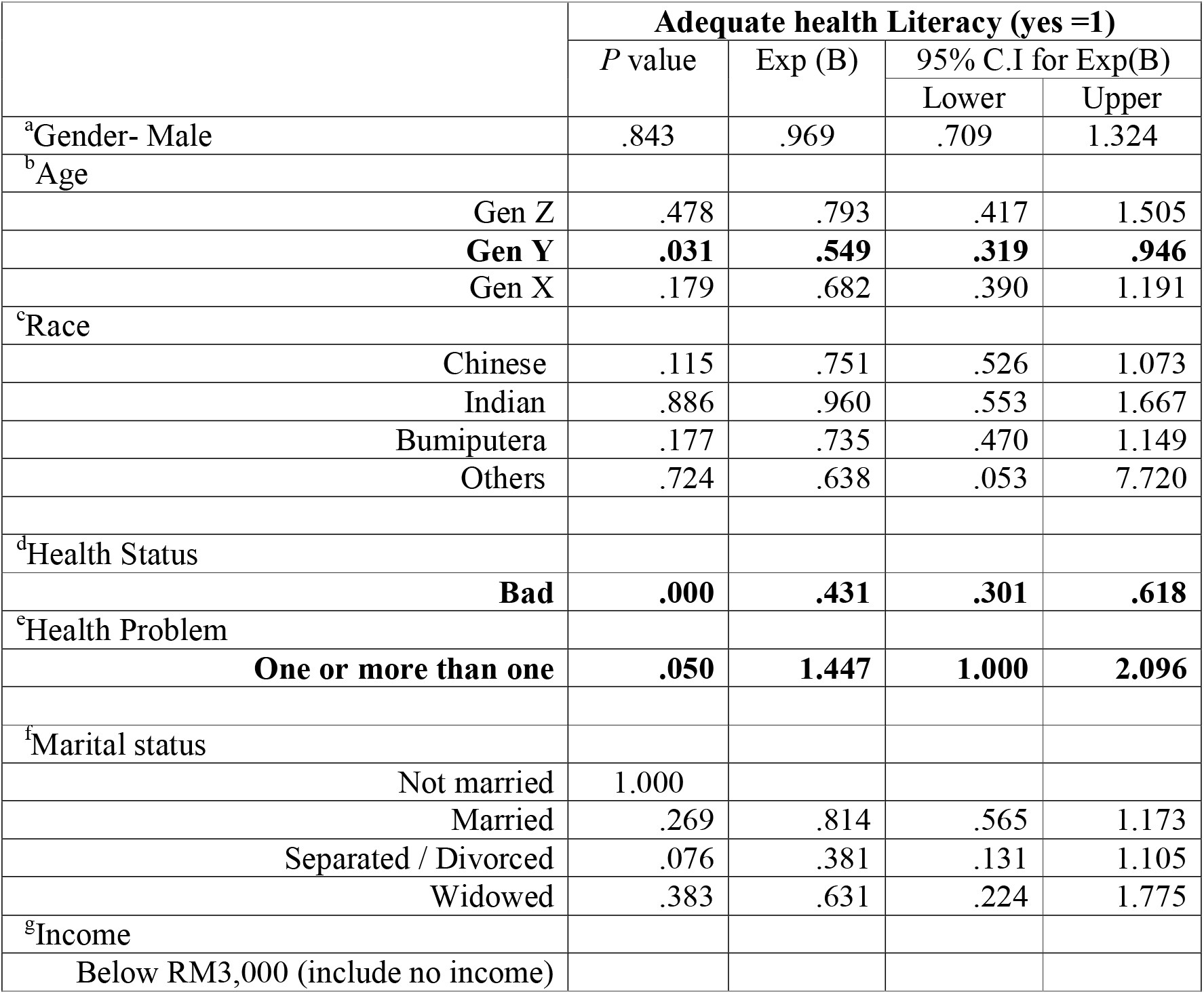
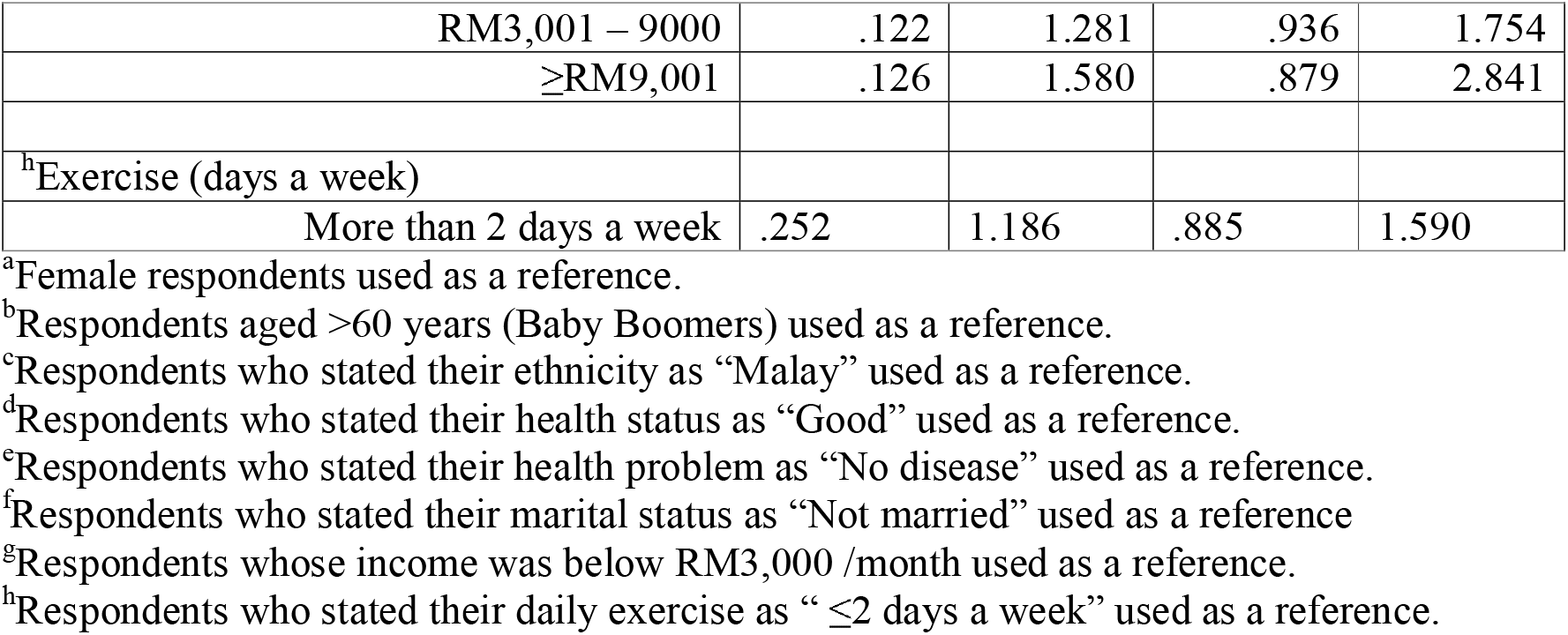
Odds ratios (95% confidence intervals) of having limited health literacy vs. adequate health literacy (N=866).

In terms of age, the findings revealed that there was a significant relationship between age and health literacy. Generation Y participants (aged 23-37) were less likely to be associated with adequate health literacy (expected beta less than 1, C.I = 0.319 - 0.946, P= 0.031 < p=0.05). Respondents who had perceived bad health status were less likely to be associated with adequate health literacy (expected beta less than 1, C.I = 0.301 - 0.618, P= 0.000 < p=0.05) compared to those who rated their health as good. This indicates that if the level of perceived bad health status increases, the odds of being associated with adequate health literacy will decrease. The association of health problems with the level of health literacy was statistically significant. Respondents who had reported that they have ‘One or more than one’ were nearly 1.5 more likely to be associated with adequate health literacy compared to those who had no disease (expected beta more than 1, C.I = 1.000 - 2.096, P= 0.05 < p=0.05). The logistic regression results also showed that the other characteristics such as gender, race, marital status, income and daily exercise remained not significantly associated with health literacy level.

## 4. Discussion

The results of our study indicate that Malaysians with one or more diseases were significantly more likely to have higher health literacy levels. The same pattern was observed in a study conducted among university students in Turkey; health literacy was significantly higher in those with chronic conditions [23]. A possible explanation for this is that people with diagnosis of long-term illness(es) were better acquainted with the healthcare system, health advice and information. However, this raises concerns regarding the point at which people begin to build higher levels of health literacy. Familiarity with health information and services as a result of a long-term illness diagnosis does not benefit the individual in terms of disease prevention, early detection and early treatment.

On the other hand, the results also showed that people with no long-term diseases were less likely to have adequate health literacy. This suggests that those with no long-term illness may be more complacent in health knowledge and behaviour, while those with long-term illness were more motivated to learn and engage in self health management. Previous studies have found similar results; individuals who were active in maintaining and improving their health were those who had higher motivation to do so [24].

In terms of health status, our findings reveal that people who perceived themselves to have bad health were less likely to have adequate health literacy. This is consistent with extant literature indicating that those with low self-rated health tend to believe that it is due to insufficient information in managing their health. The study further suggested that this led to low confidence in navigating the healthcare system thus affecting health literacy [25]. Another study found that individuals with poor or very poor self-assessment of health were more likely to have lower levels of health literacy [26].

The present study also found that people between the ages of 23 and 27 were less likely to have adequate health literacy. Research conducted in Denmark found similar results where lower health literacy was recorded among the younger population [26]. In another study, adults aged 25 to 45 years were also found to have more difficulties with health literacy compared to older individuals [27]. This is worrying considering rampant health misinformation on the internet and its widespread use among the young generation. In previous studies, millennials were found to refer to online reviews prior to deciding on a physician for consultation [28]. With negative evidence that social media contributes to the propagation of misinformation [29], this poses a threat to public health systems where accuracy of health-related information is concerned. In Malaysia, studies on eHealth literacy are still in their infancy [30, 31].

## 5. Limitations

A multi-stage sampling procedure was conducted to select the respondents in this survey. The sampling procedure prioritised ethnic group and urban/rural strata, important components in sampling multiracial populations to ensure inclusivity [16]. As a result, the gender and age distribution of the sample does not accurately reflect the current Malaysian population. The respondents of the study consisted of 65% women while current Malaysian population estimates show only 49% of the population are women. Similarly, 51% of the study sample were aged between 25 to 42 years of age; Malaysian population estimates show that only 32.9% of Malaysians are between the ages of 25 to 42 years.

The instrument utilised in this survey was the HLS-M-Q18, the shortened version of HLS-EU-Q47 tested for the Malaysian population. While this is beneficial for the overall assessment of health literacy, this has limitations in that the three health literacy domains were not measured independently.

## 6. Conclusions

Prior to the development of the Malaysian adaptation of the HLS-EU-Q47, health literacy in Malaysia was assessed utlising different instruments ranging from Newest Vital Signs [32] to tools addressing specific disease literacy such as in dentistry [33,34] and mental health [35]. The HLS-M-Q18 has enabled the measurement of health literacy in line with current global standards. Our study found that perceived health status and health problems were associated with health literacy levels. Markedly, lower health literacy levels were found to be associated with the younger generation. This is especially concerning considering this generation’s wide use of the internet as a source of information. The findings of this study have provided baseline data of the health literacy of Malaysians and provide evidence toward potential areas of intervention.

## Data Availability

The authors do not have permission to release the data. However, the data are available from The Institute for Health Behavioural Research, Ministry of Health, Malaysia upon application.

## Acknowledgments

The authors would like to thank the enumerators from the Ministry of Health Malaysia for their assistance in the data collection process. A special thanks is also extended to Ms. Suraiya Syed Mohamed, Dr. Affendi Isa and Ms. P. Komathi for their invaluable contribution to the study.

## References

1. Liu L, Qian X, Chen Z, He T. Health literacy and its effect on chronic disease prevention: evidence from China’s data. BMC Public Health. 2020;20:690.

2. Gonza’lez-Chica DA, Mnisi Z, Avery J, Duszynski K, Doust J, Tideman P, et al. Effect of health literacy on quality of life amongst patients with ischaemic heart disease in Australian general practice. PLoS One. 2016;11:e0151079.

3. Santos P, Sá L, Couto L, Hespanhol A. Health literacy as a key for effective preventive medicine, Cogent Soc Sci. 2017;3(1):1407522.

4. Jandorf S, Nielsen MK, Sørensen K, Sørensen TL. Low health literacy levels in patients with chronic retinal disease. BMC Ophthalmol. 2019;19(1):174.

5. Anwar WA, Mostafa NS, Hakim SA, Sos DG, Abozaid DA, Osborne RH. Health literacy strengths and limitations among rural fishing communities in Egypt using the Health Literacy Questionnaire (HLQ). PLoS ONE. 2020;15(7):p e0235550.

6. McCaffery KJ, Dodd RH, Cvejic E, Ayre J, Batcup C, Isautier JMJ, et al. Health literacy and disparities in COVID-19–related knowledge, attitudes, beliefs and behaviours in Australia. Public Health Res Pract. 2020;30(4):e30342012.

7. Haun JN, Patel NR, French DD, Campbell RR, Bradham DD, Lapcevic WA. Association between health literacy and medical care costs in an integrated healthcare system: a regional population based study. BMC health Serv Res 2015;15:249.

8. Lopes-Marques SR, Aguiar-Lemos, SM. Health literacy assessment instruments: literature review. Audiology Communication Research. 2017;22. e1757

9. Guo S, Armstrong R, Waters E, Sathish T, Alif SM, Browne GR, Yu X. Quality of health literacy instruments used in children and adolescents: a systematic review. BMJ Open 2018;8:e020080.

10. Abdullah A, Liew SM, Salim HS, Ng CJ, Chinna K. Health literacy research in Malaysia: A scoping review. Sains Malaysiana. 2020;49:5.

11. Duong TV, Aringazina A, Baisunova G, Nurjannah PTV, Pham KM, Truong TQ, et al. Measuring health literacy in Asia: validation of the HLS-EU-Q47 survey tool in six Asian countries. J Epidemiol. 2017;27(2):80–6.

12. Mohamad EMW, Kaudan MK, Hamzah MR, Azlan AA. Ayub SH, Tham, JS, et al. Establishing the HLS-M-Q18 short version of the European health literacy survey questionnaire for the Malaysian context. BMC Public Health. 2020;20:580.

13. Dodson S, Good S, Osborne RH. Health literacy toolkit for low- and middle-income countries: a series of information sheets to empower communities and strengthen health systems. New Delhi: 2015.World Health Organization, Regional Office for South-East Asia.

14. Trezona A, Dodson S, Osborne RH. Development of the Organizational Health Literacy Responsiveness (Org-HLR) Framework in Collaboration with Health and Social Services Professionals. BMC Health Services Research. 2017;17:513. pmid:28764699

15. Froze S, Arif MT, Saimon R. Does health literacy predict preventive lifestyle on metabolic syndrome? A population-based study in Sarawak Malaysia. Open J Prev Med. 2018;8(6):169.

16. Charmaraman L, Woo M, Quach A, Erkut S. How have researchers studied multiracial populations? A content and methodological review of 20 years of research. Cult Divers Ethn Minor Psychol. 2014;20(3):336–52.

17. Berens EM, Vogt D, Ganahl K, Weishaar H, Pelikan J, Schaeffer D. Health literacy and health service use in Germany. Health Lit Res Pract. 2018;2(2):115–22.

18. Ministry of Rural Development. The Rural Master Plan. Putrajaya: Kementerian Kemajuan Luar Bandar dan Wilayah; 2010.

19. Koran J. Preliminary proactive sample size determination for confirmatory factor analysis models. Meas Eval Couns Dev. 2017;49(4):296–308.

20. Conroy R. Sample size: a rough guide [Internet]. Ethics (Medical Research) Committee; 2015 [cited 2021 Jan 14]. Available from: http://www.beaumontethics.ie/docs/application/samplesizecalculation.pdf.

21. Israel GD. Determining sample size. Gainesville: University of Florida; 1992. Report No.:Fact Sheet PEOD-6.

22. Brislin RW. Back-translation for cross-cultural research. J Cross Cult Psychol. 1970; 3(1): 185–216.

23. Uysal N, Ceylan E, Koç A. Health literacy level and influencing factors in university students. Health & Social Care in the Community. 2019:hsc.12883.

24. Smith SG, Curtis LM, Wardle J, von Wagner C, Wolf MS. (2013). Skill set or mind set? Associations between health literacy, patient activation and health. PLoS One. 2013;8(9):e74373.

25. Storey A, Hanna L, Missen K, Hakman N, Osborne RH, Beauchamp A. The association between health literacy and self-rated health amongst Australian university students. J Health Commun. 2020;25(4):333–343.

26. Svendsen MT, Bak CK, Sørensen K, Pelikan J, Riddersholm SJ, Skals RK, et al. Associations of health literacy with socioeconomic position, health risk behavior, and health status: a large national population-based survey among Danish adults. BMC Public Health. 2020;20:1–12.

27. Bo A, Friis K, Osborne RH, Maindal HT. National indicators of health literacy: ability to understand health information and to engage actively with healthcare providers - a population-based survey among Danish adults. BMC Public Health. 2014;14:1095.

28. Rohrich RJ, Dayan E. Improving communication with millennial patients. Plast Reconstr Surg. 2019;144(2):533–535.

29. McKee M, van Schalkwyk MCI, Stuckler D. The second information revolution: digitalization brings opportunities and concerns for public health. Eur J Environ Public Health. 2019;29:3–6

30. Azlan AA. Measures of eHealth literacy: options for the Malaysian population. J Komun Malays J Commun. 2019;35(4):211–228.

31. Yilma TM, Inthiran A, Reidpath DD, Orimaye SO. Context-based interactive health information searching. Information Research. 2019;24(2):1–22.

32. Norrafizah J, Nor Asiah M, Suraiya SM, Zawaha HI, Normawati A. Assessment of health literacy among people in a rural area in Malaysia using newest vital signs assessment. Journal of Education, Society and Behavioural Science. 2016;16(2):1–7.

33. Rani H, Su YH, Ding SA, Yahya NA, Jaafar A. Comparison of visual oral health literacy level pre and post oral health education among adolescents. Journal of International Dental and Medical Research. 2019;12(2):640–644.

34. Muhd Noor N, Rani H, Zakaria ASI, Yahya NA, Sockalingam SN. Sociodemography, oral health status and behaviours related to oral health literacy. Pesqui Bras Odontopediatria Clin Integr. 2019;19(1):1–10.

35. Ibrahim N, Amit N, Shahar S, Wee LH, Ismail R, Khairuddin R, et al. Do depression literacy, mental illness beliefs & stigma influence mental health help-seeking attitude? A cross-sectional study of secondary school and university students from B40 households in Malaysia. BMC Public Health. 2019; 1-8:544.

